# Investigating the causal relationship between physical activity and chronic back pain: a bidirectional two-sample Mendelian randomization study

**DOI:** 10.1101/2021.07.20.21260847

**Authors:** Shaowei Gao, Huaqiang Zhou, Siyu Luo, Xiaoying Cai, Fang Ye, Qiulan He, Chanyan Huang, Xiaoyang Zheng, Ying Li, Zhanxin Du, Yaqing Wang, Zhihui Qi, Zhongxing Wang

## Abstract

**Background:** Recent observational studies have reported a negative association between physical activity and chronic back pain (CBP), but the causality of the association remains unknown. We introduce bidirectional Mendelian randomization (MR) to assess potential causal inference between physical activity and CBP.

**Methods:** The two-sample MR was used with independent genetic variants associated with physical activity phenotypes and CBP as genetic instruments from large genome-wide association studies (GWASs) on individuals of European ancestry. The effects of both directions (physical activity to CBP and CBP to physical activity) were examined. Inverse variance-weighted meta-analysis and alternate methods (weighted median and MR-Egger) were used to combine the MR estimates of the genetic instruments. Multiple sensitivity analyses were conducted to examine the robustness of the results.

**Results:** For primary analysis, instrumental variables were extracted from 337,234 participants for physical activity (the same as the outcome cohort) and 158,025 participants (29,531 cases) for CBP, while the outcome cohort for CBP included 117,404 participants (80,588 cases). No evidence of a causal relationship was found in the direction of physical activity to CBP (odds ratio [OR], 0.98; 95% CI, 0.85-1.13; P = 0.81). In contrast, a negative causal relationship in the direction of CBP to physical activity was detected (β = -0.07; 95% CI, -0.12 to -0.01; P = 0.02), implying a reduction in moderate-vigorous physical activity (approximately 146 MET-minutes/week) for participants with CBP relative to controls.

**Conclusions:** The negative relationship between physical activity and CBP is probably derived from the reduced physical activity of patients experiencing CBP rather than the protective effect of physical activity on CBP.

**Highlights:**

➢ Previous studies found a negative relationship between physical activity and chronic back pain, but the causal inference behind the relationship is lacking in evidences.
➢ We applied Mendelian randomization and revealed that the negative relationship probably derived from the fact that patients experiencing CBP tend to reduce their physical activities.
➢ If the negative relationship between physical activity and CBP is truly a reverse causality, the concept that patients with CBP should be engaging in activity, which is recommended by current guidelines, may need to be reconsidered.

## 1 Introduction

Back pain, especially low back pain, has become a large burden worldwide, as it is estimated to affect more than 510 million people and cause over 57 million “years lived with disability” in 2016^1^. At least one-third of patients with back pain report persistent pain after an acute episode and eventually develop chronic back pain (CBP)^2^, which is generally defined as back pain lasting ≥ 3 months^3^. A key step in preventing CBP is the identification of possible risk factors, especially intervenable risk factors. To date, well-known risk factors for CBP have included smoking^4^, obesity^5^, previous episodes of back pain^6^, other chronic conditions (e.g., diabetes, headache)^7^, and poor mental health^8, 9^. However, the role of physical activity on CBP is inconclusive (Table 1).

**Table 1.**
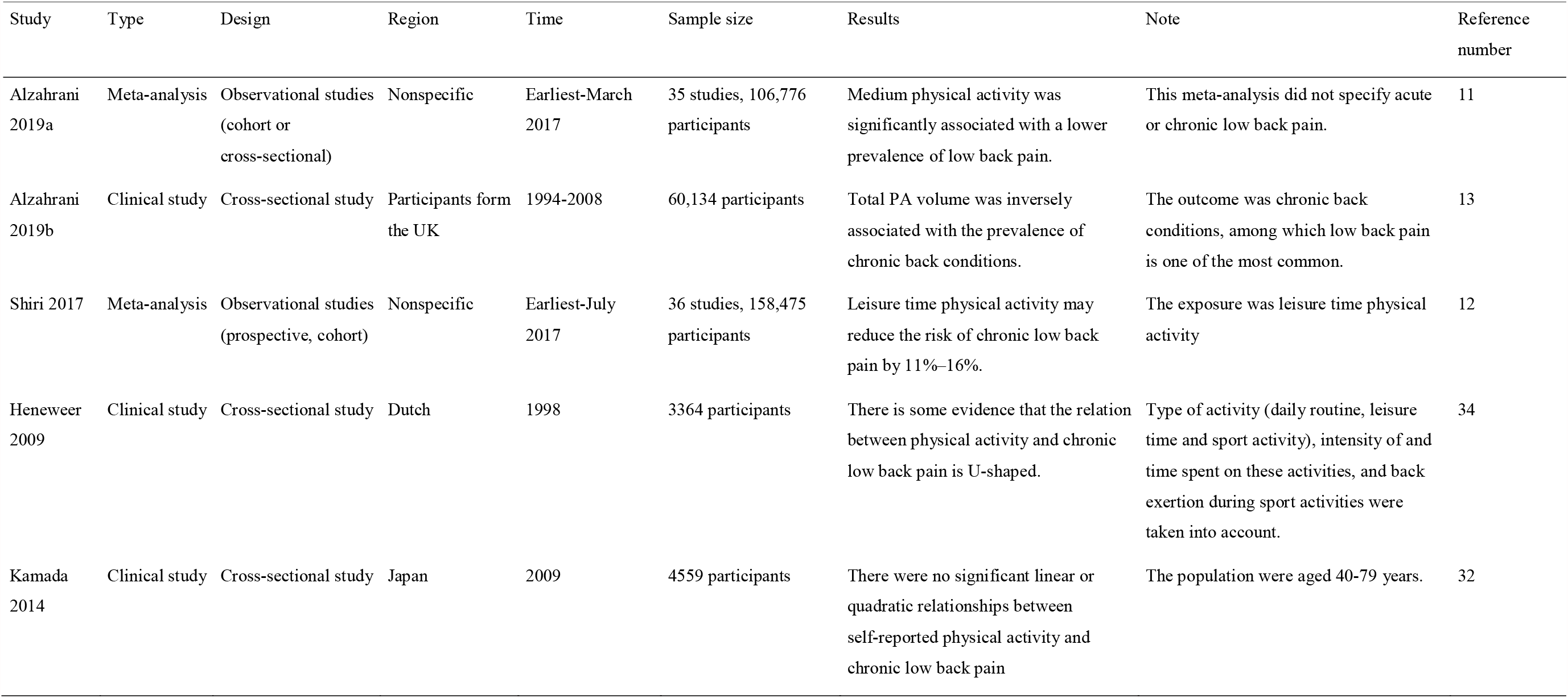
Representative studies for the association between physical activity and CBP

| Study | Type | Design | Region | Time | Sample size | Results | Note | Reference number |
| --- | --- | --- | --- | --- | --- | --- | --- | --- |
| Alzahrani 2019a | Meta-analysis | Observational studies (cohort or cross-sectional) | Nonspecific | Earliest-March 2017 | 35 studies, 106,776 participants | Medium physical activity was significantly associated with a lower prevalence of low back pain. | This meta-analysis did not specify acute or chronic low back pain. | 11 |
| Alzahrani 2019b | Clinical study | Cross-sectional study | Participants from the UK | 1994-2008 | 60,134 participants | Total PA volume was inversely associated with the prevalence of chronic back conditions. | The outcome was chronic back conditions, among which low back pain is one of the most common. | 13 |
| Shiri 2017 | Meta-analysis | Observational studies (prospective, cohort) | Nonspecific | Earliest-July 2017 | 36 studies, 158,475 participants | Leisure time physical activity may reduce the risk of chronic low back pain by 11%–16%. | The exposure was leisure time physical activity | 12 |
| Heneweer 2009 | Clinical study | Cross-sectional study | Dutch | 1998 | 3364 participants | There is some evidence that the relation between physical activity and chronic low back pain is U-shaped. | Type of activity (daily routine, leisure time and sport activity), intensity of and time spent on these activities, and back exertion during sport activities were taken into account. | 34 |
| Kamada 2014 | Clinical study | Cross-sectional study | Japan | 2009 | 4559 participants | There were no significant linear or quadratic relationships between self-reported physical activity and chronic low back pain | The population were aged 40-79 years. | 32 |

Physical activity is defined as musculoskeletal movement that results in energy consumption^10^. As shown in Table 1, recent meta-analyses reviewed tens of observational studies and found a negative relationship between physical activity and CBP^11, 12^. A similar conclusion was also reported by other cross-sectional studies^13, 14^. However, studies with high-level evidence (such as randomized control studies), which can address the problem of causal inference, are lacking. Consequently, whether the negative relationship between physical activity and CBP is due to the protective effect of physical activity on CBP or the tendency of patients with CBP to reduce physical activity remains unknown.

Randomized control studies on physical activity are difficult to conduct, as it is unethical to constrain participants’ physical activity. Mendelian randomization (MR) is an alternative method to achieve randomization for this situation by treating genetic variation as a natural experiment in which individuals are randomly assigned to different levels of nongenetic exposure during their lifetime^15^. In addition, MR can strengthen causal inferences by importing a bidirectional design.

In this study, we first applied bidirectional MR to explore the association between physical activity and CBP (Figure 1). We aim to clarify the causal inference behind this association. We hypothesize that physical activity has inverse causal association with chronic back pain.

**Figure 1.**
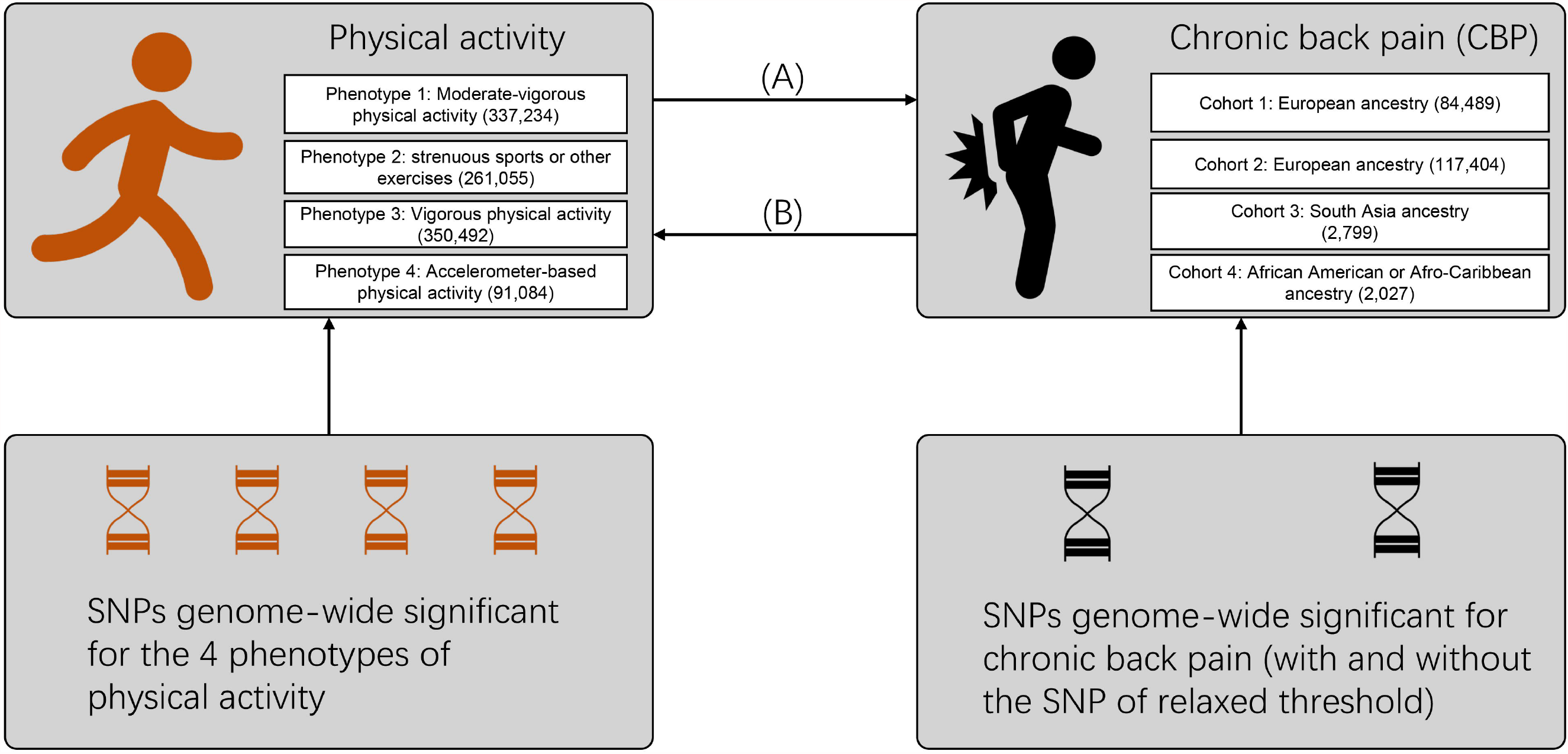
Flow diagram for the design of the bidirectional, two-sample Mendelian randomization study. Multiple phenotypes and cohorts were cross-validated to maintain the robustness of our results. The direction marked (A) refers to the effect of physical activity on chronic back pain, while that marked (B) refers to the reverse effect. Details on the SNPs used as trait instruments are summarized in Tables S2-S4. The numbers of participants for different phenotypes or cohorts are labeled in the brackets. SNP: single-nucleotide polymorphism.

## 2 Methods

This is a Mendelian randomization study with a bidirectional and two-sample design, as illustrated in Figure 1. All the data used are summary-level and derived from public genome-wide association studies (GWAS), which had obtained ethical permissions from their respective institutional review boards and written informed consent from their respective participants. **Patients and public were not involved in this MR study**. The study was conducted under Burgess’s guidelines (Supplementary checklist 1)^16^ and reported according to the STROBE-MR statement (Supplementary checklist 2)^17^. We analyzed these data from April 20, 2021 to June 20, 2021.

### 2.1 Selection of instruments and outcome data

#### 2.1.1 Physical activity

The physical activity instruments were based on Klimentidis’s GWAS conducted with participants of the UK Biobank cohort^18^. This GWAS, using a population of predominantly European ancestry, examined the following four physical activity phenotypes: (1) self-reported moderate-vigorous physical activity (continuous phenotype, 337,234 participants, in standardized units of inverse normalized metabolic equivalent minutes per week [MET-minutes/week]), (2) self-reported vigorous physical activity (binary phenotype, 261,055 participants with 98,060 cases, ≥ 3 versus 0 for days per week), (3) self-reported strenuous sports or other exercises (binary phenotype, 350,492 participants with 124,842 cases, ≥ 2-3 versus 0 for days per week), and (4) seven-day average acceleration from a wrist-worn accelerometer (continuous phenotype, 91,084 participants, in milligravities). The characteristics for each phenotype are summarized in Table S1. We chose single-nucleotide polymorphisms (SNPs) from the first phenotype (self-reported moderate-vigorous physical activity) for the primary analysis, as this phenotype yielded the largest number of significant SNPs. To ensure robustness, the SNPs from the other three phenotypes were used in a sensitivity analysis (Table S2). In addition, as the GWAS of the accelerometer-based activity identified only 2 SNPs but had higher heritability than that of the self-reported activity (∼14% vs ∼5%), the top SNPs meeting a relaxed threshold (P < 1×10^−7^) were also imported to our study (Table S3) in a sensitivity analysis; the method of using SNPs with relaxed thresholds has been used for other MR studies when insufficient SNPs are available^19-21^. We retained only the top independent SNPs by selecting one representative SNP among highly correlated SNPs (r^2^ > 0.001), a process known as ‘clumping’. If an instrument SNP was not present in the outcome GWAS, then a proxy SNP that was in linkage disequilibrium with the instrument SNPs was searched for instead. Clumping and proxy SNPs are both based on reference data from the 1000 Genomes Project^22^.

For the other direction, in which physical activity is regarded as the outcome trait, we again applied Klimentidis’s GWAS^18^. The completed summary data can be accessed from the OpenGWAS database through the MR-base platform^23, 24^. Similarly, data for all 4 phenotypes above are available, while moderate-vigorous physical activity was used for the primary analysis.

#### 2.1.2 Chronic back pain

Genetic instruments for CBP were derived from a genome-wide meta-analysis comprising adults of European ancestry from 16 cohorts^25^, in which positive cases were obtained by examining the questionnaires from the participants. These cohorts did not have a consistent definition of CBP: two cohorts used “≥ 1 month of back pain in consecutive years”; nine cohorts used “≥ 6 months of back pain”; six cohorts used “≥ 3 months of back pain”. The control group enrolled participants who reported not having back pain or reported back pain of insufficient duration as cases. Most of the included cohorts did not include question items regarding localization of the pain to the low back or lumbar region specifically. Therefore, a general definition examining chronic ‘back pain’ rather than a more specific chronic ‘low back pain’ definition was applied. This meta-analysis identified 4 SNPs tightly related to chronic back pain, one of which met a relaxed threshold (P = 3.9×10^−7^), while the others met strict criteria (P < 5×10-8) (Table S4). Similarly, we introduced a sensitivity analysis by eliminating the SNP with a relaxed threshold.

For the outcome data, we searched the OpenGWAS database and found 4 GWAS cohorts with completed summary data (Table S5). Two out of the four cohorts are of European ancestry, while the other two contain South Asian populations and African American or Afro-Caribbean populations. Because the MR results may be uninformative for the magnitude (rather than the direction) of the effect when the exposure and outcome studies are derived from different populations^24^, we selected one European ancestry cohort with the maximum sample size (117,404 participants and 80,588 cases) for the primary analysis and the other three for the sensitivity analyses.

### 2.2 Statistical analysis

The R package “TwoSampleMR” developed by researchers in the MR-base platform was used for this Mendelian randomization study^24^. Briefly, the algorithm in this package combines the effect sizes of the instruments on exposure traits with those of the instruments on outcome traits using the principle of meta-analysis. In addition to the effect size, the effect allele and its frequency for each instrument — whether for exposure or outcome — must be extracted to determine the direction of the strand.

As the primary method for combining MR estimates, we used the multiplicative random-effect inverse variance-weighted (IVW) method, which translates to a weight regression of instrument-outcome effects on instrument-exposure effects where the intercept is restricted to zero^26^. In this way, bias may occur if horizontal pleiotropy (in which the instruments influence the outcome through causal pathways other than the exposure) is present. We therefore introduced two other MR methods: the weighted median method and MR-Egger regression. The weighted median method chooses the median MR estimate of the instruments as the result, while MR-Egger regression allows the intercept to be a value other than zero^27, 28^. Both methods are more robust for horizontal pleiotropy, although at the cost of reduced statistical power^29^. Generally, the effect size for the binary outcome should be represented as odds ratio (OR) (i.e., exponentiated β). However, in Klimentidis’s GWAS, a mixed model-model linear regression was used even for binary phenotypes (vigorous PA and strenuous sports or other exercises), leading to unreliable estimates of effect sizes (but not influencing the direction and statistical power)^18^. We therefore reported the effect estimates in the β value for PA as an outcome trait (we avoided translating the meaning of β for the binary phenotypes) and in the OR for CBP as an outcome trait.

A series of methods were applied for the sensitivity analyses: in addition to setting multiple comparisons among different phenotypes and different cohorts, the funnel plot, Cochran’s Q statistic, leave-one-out analyses, MR-Pleiotropy RESidual Sum and Outlier (MR-PRESSO), and the MR-Egger intercept test of deviation from the null were used to detect heterogeneity and horizontal pleiotropy^30^. By implementing a homonymous R package, MR-PRESSO also detects and corrects outlier SNPs reflecting pleiotropic biases^31^. Finally, to determine potential pleiotropy, we searched each instrument used for the primary analysis in the PhenoScanner GWAS database (version 2; http://phenoscanner.medschl.cam.ac.uk) to find any existing associations with potential confounding traits; then, we removed these SNPs to control the pleiotropic effects and to see if the primary results could be reversed.

## 3 Results

### 3.1 Effect of physical activity on CBP

In this direction, we found no evidence of a causal relationship between physical activity and CBP. In our primary analysis — the effect of self-reported moderate-vigorous physical activity on the largest CBP cohort with European ancestry — the combined IVW OR was close to 1 (IVW OR, 0.98; 95% CI, 0.85-1.13; P = 0.81) (Table 2, Figure 2), which supported acceptance of the null hypothesis that there is no effect of physical activity on CBP. The results were almost consistent for different exposure phenotypes and different outcome cohorts (Table S6). The funnel plot did not detect obvious asymmetry, and the leave-one-out analysis did not change the pattern of the result (Figure S1). The MR-Egger intercept test suggested no directional horizontal pleiotropy (intercept, 0.001; standard error, 0.005; P = 0.81), even though Cochran’s Q test indicated moderate heterogeneity (Q = 19.8; P = 0.011). MR-PRESSO detected one outlier (rs1043595), but the result remained negative when this outlier was removed (Table 2).

**Table 2.** MR results for the effect of self-reported moderate-vigorous physical activity on CBP

| Method | OR (95% CI)<sup>b</sup> | P Value | No. of SNPs |
| --- | --- | --- | --- |
| With outlier<sup>a</sup> |  |  |  |
| IVW | 0.98 (0.85-1.13) | 0.81 | 9 |
| Weighted median | 0.96 (0.84-1.11) | 0.59 | 9 |
| MR-Egger | 0.91 (0.48-1.73) | 0.77 | 9 |
| Without outlier<sup>a</sup> |  |  |  |
| IVW | 0.94 (0.84-1.05) | 0.26 | 8 |
| Weighted median | 0.96 (0.84-1.09) | 0.51 | 8 |
| MR-Egger | 1.00 (0.61-1.63) | 1.00 | 8 |

<sup>a</sup> the outlier was rs1043595, which was detected with the MR-PRESSO method
<sup>b</sup> indicates odds for CBP per 1-SD increase in moderate-vigorous physical activity (1-SD equals 2084 MET-minutes/week in Klimentidis's GWAS)
Abbreviations: IVW, inverse variance weighted; CBP, chronic back pain; MR, Mendelian randomization; OR, odds ratio; CI, confidence interval; SNP, single-nucleotide polymorphism

**Figure 2.**
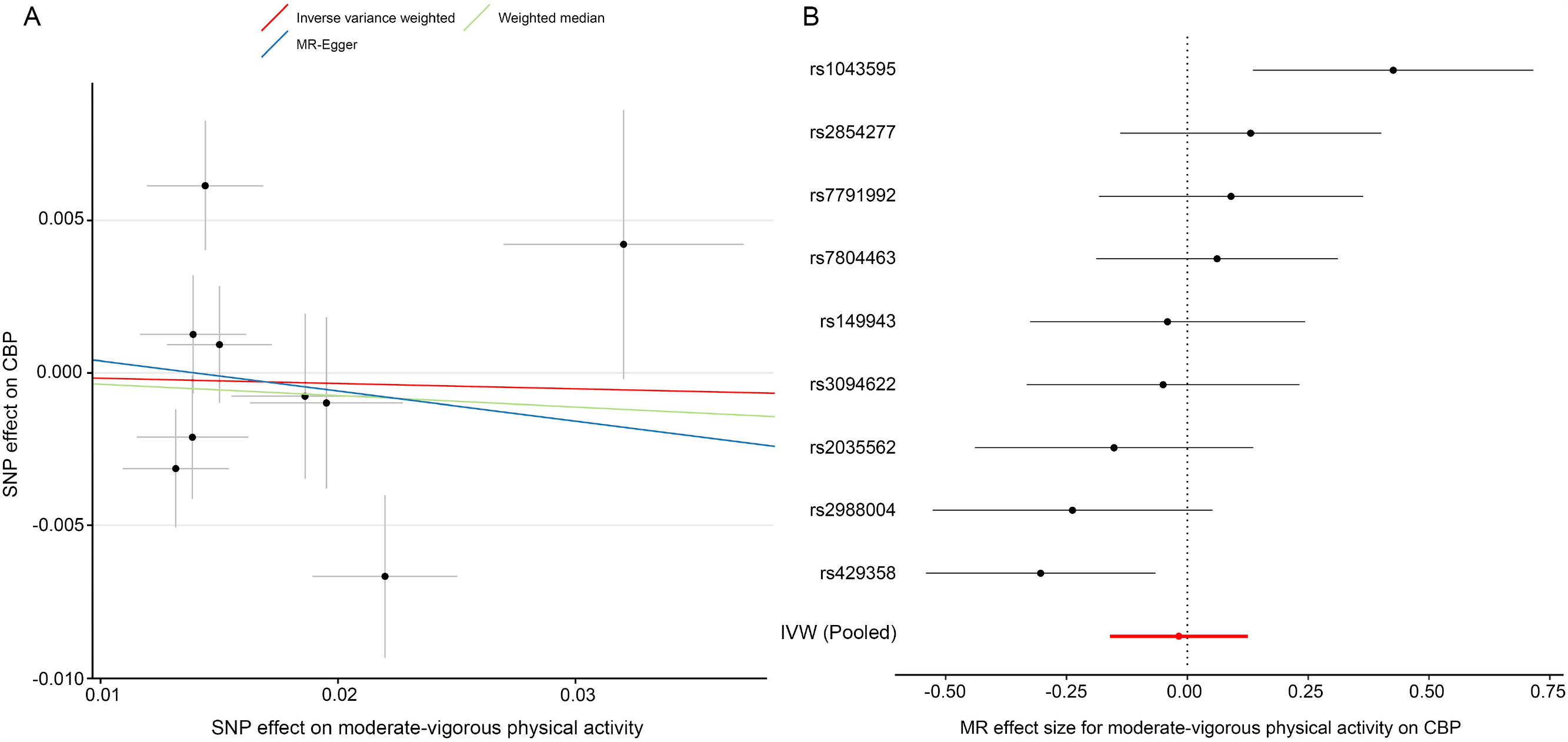
MR plots for the effect of moderate-vigorous physical activity on CBP. (A) Scatter plot of the SNP effect on moderate-vigorous physical activity vs. that on CBP. The slope of each fitted line represents the pooled MR effect calculated by each method. (B) Forest plot of individual and pooled MR effect sizes for moderate-vigorous physical activity on CBP. Each point and its corresponding line represent the β value with its 95% CI, respectively. Abbreviations: SNP, single-nucleotide polymorphism; CBP, chronic back pain; MR, Mendelian randomization; IVW, inverse variance weighted

### 3.2 Effect of CBP on physical activity

In contrast to the previous analysis, we found a robust negative causal relationship between CBP and physical activity. In our primary analysis — the effect of CBP represented by all 4 SNPs on self-reported moderate-vigorous physical activity — the MR estimate with the IVW method was significantly less than zero (IVW β, -0.07; 95% CI, -0.12 to -0.01; P = 0.02) (Table 3, Figure 3), implying that participants with CBP tended to reduce their physical activity by approximately 146 MET-minutes/week with respect to those without CBP. The weighted median and MR-Egger tests yielded similar patterns of effects (Table 3). The results were consistent not only with analyses with different outcome traits, such as self-reported strenuous sports and accelerometer-based physical activity, but also with analyses where the SNP with the relaxed threshold was removed for CBP (Table S7). The leave-one-out analysis showed that no single SNP was strong for reversely driving the overall effect of CBP on physical activity but detected one SNP (rs12310519) that played a relatively predominant role (Figure S2A). Furthermore, the funnel plot presents with a symmetric pattern (Figure S2B), and Cohran’s Q test suggested no heterogeneity (Q = 0.3; P = 0.96). In addition, MR-PRESSO found no outliers, and the MR-Egger intercept test indicated no consistent pleiotropy (intercept, 0.001; standard error, 0.004; P = 0.91).

**Table 3.** MR results for the effect of CBP on self-reported moderate-vigorous physical activity

| Method | $\beta$ (95% CI)<sup>a</sup> | P Value | No. of SNPs<sup>b</sup> |
| --- | --- | --- | --- |
| IVW | -0.07 (-0.12 to -0.01) | 0.02 | 4 |
| Weighted median | -0.07 (-0.13 to -0.01) | 0.03 | 4 |
| MR-Egger | -0.08 (-0.25 to 0.09) | 0.47 | 4 |

<sup>a</sup> indicates a change in multiple of SD of moderate-vigorous physical activity (1-SD equals 2084 MET-minutes/week in Klimentidis's GWAS) for participants with CBP vs control status
<sup>b</sup> no outlier was detected with MR-PRESSO
Abbreviations: IVW, inverse variance weighted; CBP, chronic back pain; MR, Mendelian randomization; SNP single-nucleotide polymorphism

**Figure 3.**
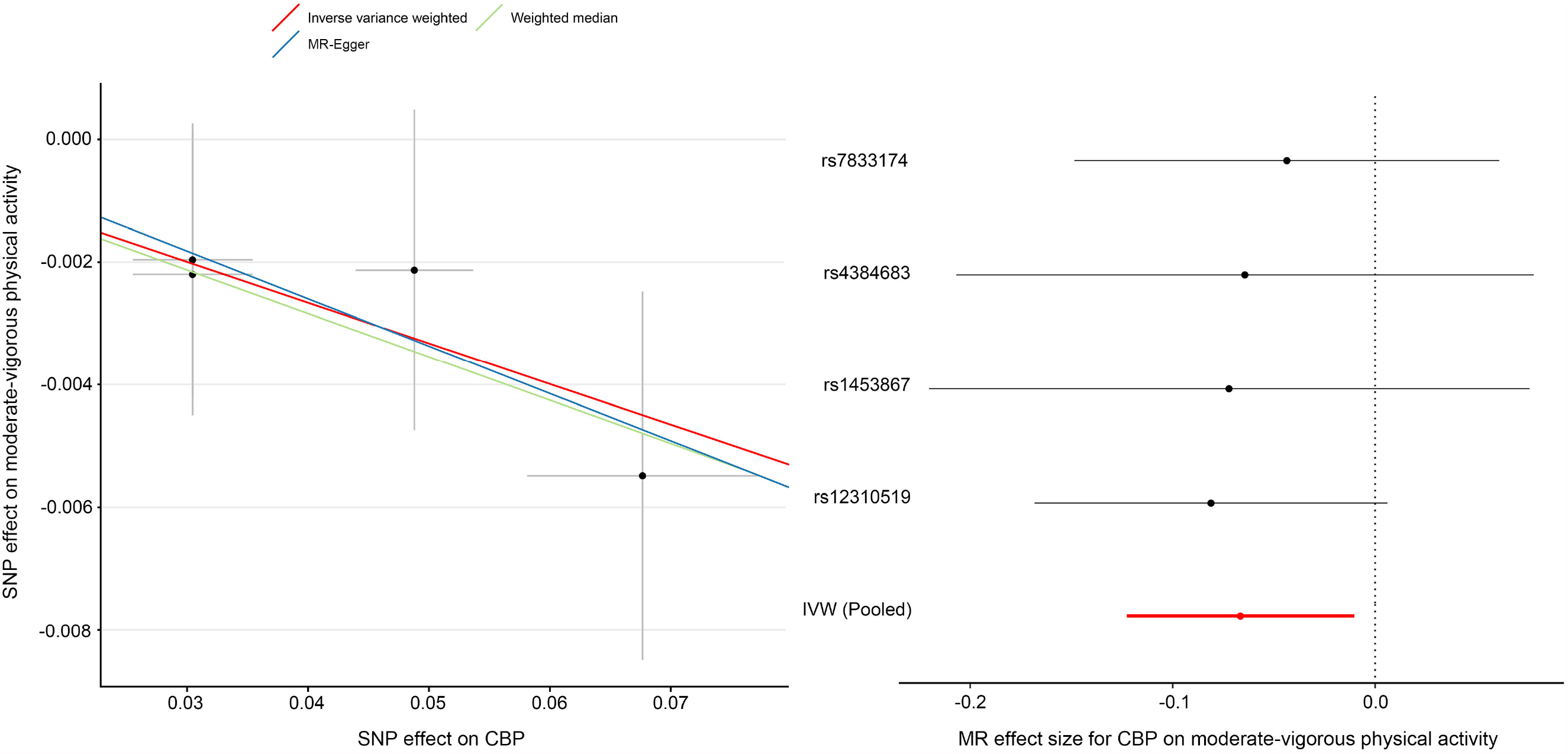
MR plots for the effect of CBP on moderate-vigorous physical activity. (A) Scatter plot of the SNP effect on CBP vs. that on moderate-vigorous physical activity. The slope of each fitted line represents the pooled MR effect calculated by each method. (B) Forest plot of individual and pooled MR effect sizes for CBP on moderate-vigorous physical activity. Each point and its corresponding line represent the β value and its 95% CI, respectively. Abbreviations: SNP, single-nucleotide polymorphism; CBP, chronic back pain; MR, Mendelian randomization; IVW, inverse variance weighted

### 3.3 Potential pleiotropy searched in PhenoScanner

In total, thirteen SNPs were included in our primary analyses (9 for physical activity to CBP, 4 for CBP to physical activity). The most potential pleiotropy was “trunk fat/fat-free mass”, which was involved in 7/13 of all SNPs (Table S8). After removing these SNPs, the pattern of the primary results did not change (physical activity to CBP: IVW OR, 1.09; 95% CI 0.85-1.40; P = 0.52; CBP to physical activity: IVW β, -0.077; 95% CI, -0.15 to -0.003; P = 0.04) (Figure S3).

## 4 Discussion

To the best of our knowledge, this is the first MR study to explore the causal relationship between physical activity and CBP. We examined the effects in both directions and found that engaging in more physical activity was not associated with a reduced risk of CBP, but having CBP was associated with reduced physical activity (including both self-reported and accelerometer-based physical activity). The result supports the more intuitive view that the negative association between physical activity and CBP arises from the fact that patients with CBP tend to reduce their physical activity.

### 4.1 Comparisons with previous traditional studies

Previous studies reported conflicting results regarding the effect of physical activity on CBP. Some studies showed no association between physical activity and CBP^32, 33^ or a U-shaped relationship, in which very low and very high levels of physical activity increased the risk of CBP^34^. However, a recent observational study with a large population and two meta-analyses supported a negative relationship between physical activity and CBP^11-13^. The observational study involved a population 60,134 adults, but its cross-sectional design was insufficient for identifying the causal inference between physical activity and CBP^13^. Although the meta-analysis recruited prospective studies^12^, the observational design was “apt in generating hypotheses and suggesting causality but can never prove it”^35^. In contrast, MR can mimic the design of randomized controlled trials^24^. Given that a an SNP is known to be related to a trait (the so-called “instrument variable”), according to Mendel’s law, the alleles at the SNP are causally upstream of the corresponding trait and expected to be random with respect to potential confounders. In an MR study, participants are randomly assigned to the treatment group or control group according to the genotype at the instrument SNP of exposure. Then, the effect size of the causal inference can be calculated as the ratio between the SNP effect on the outcome and the SNP effect on the exposure. Our study extends the current literature from the level of association to the level of causal inference.

### 4.2 Robustness

Our results were robust to different pairs of exposure and outcome cohorts (Table S6 & S7). In the direction of physical activity to CBP, engaging in more physical activity did not significantly change the risk of CBP except in the “ukb-e-3571_AFR” cohort (Table S6). The small sample size (approximately 2000 participants) of the “ukb-e-3571_AFR” cohort and the wide range of the OR indicate that the exception probably derives from a random error. In the other direction, from CBP to physical activity, reporting CBP was always associated with reporting reduced physical activity (Table S7). However, in the leave-one-out analysis, we found one predominant SNP, rs12310519, without which the OR of reporting CBP on reporting moderate-vigorous physical activity was no longer statistically significant (the 95% CI for the OR included 1) (Figure S2A). To examine the extent of the influence of this SNP, we repeated the leave-one-out analysis on the other three phenotypes of physical activity; interestingly, however, this SNP (rs12310519) was not the predominant SNP for self-reported vigorous physical activity and self-reported strenuous sports or other exercises (Figure S4). This result may imply different mechanisms by which genetic variance influences different levels of one phenotype.

After looking up the SNPs used for the primary analysis in the Phenoscanner database, a potential pleiotropy, “trunk fat/fat-free mass”, was detected. This trait has been reported as a common predictive factor for both physical activity and CBP^36-38^ and served as an exposure-outcome confounder for the current study. Nevertheless, the pattern of the primary results did not change after controlling this pleiotropy, possibly due to the balance of the multiple SNPs that have effects of different directions on this confounder.

### 4.3 Limitations

This study has several limitations. First, although different levels of physical activity were included in this study, the CBP was an all-or-none variable. Thus, it was impossible to compare the effect between different levels of CBP. Second, there were overlapping samples in both the exposure and outcome studies because the physical activity source study and the CBP outcome data both involved participants from the UK Biobank project. Results from MRs with overlapping samples may be biased due to the winner’s curse phenomenon^39^. However, we used a sensitivity analysis in which weaker instruments were excluded, which can minimize the bias from sample overlap^40^. Finally, the CBP phenotype we used represents a symptom rather than a disease or a biomarker. Compared with other more detailed phenotypes, such as osteoarthritis, additional mechanisms may be involved in CBP, such as muscle injury, nerve root compression, or intervertebral disc degeneration. Thus, a single genome-wide association study is insufficient for finding all SNPs as instruments for CBP. Although the genome-wide meta-analysis we selected for this MR included 16 CBP cohorts, it detected only three to four SNPs, which might partially cover all the mechanisms.

Another point we should clarify is that we used chronic back pain instead of chronic low back pain, a more commonly used phenotype, as the exposure phenotype. The primary reason for this is that the questionnaires used for the included cohorts did not specifically isolate the low back region^25^. Given the high agreement between general back pain and low back pain-specific questions^41^ and since upper/mid back pain without concurrent low back pain is uncommon^42^, we believe that our results with CBP can well represent those with chronic low back pain, as exemplified in other studies using similar substitutions^25, 43^.

### 4.4 Importance

Despite these limitations, the MR study performed here provides a novel insight into genetic variants as instruments for assessing the causal inference between physical activity and CBP and obviates typical challenges in observational research while providing an internal explanation for such studies^11-13^. If the negative relationship between physical activity and CBP is truly a reverse causality, the concept that patients with CBP should be engaging in activity, which is recommended by current guidelines^44^, may need to be reconsidered.

## 5 Conclusion

This study applied MR to examine the causal inference between physical activity and CBP. The negative relationship between these two traits is probably derived from the fact that patients experiencing CBP tend to reduce their physical activities. Overall, our findings denied the protective effect of physical activity on CBP.

## Supporting information

Supplementary checklist 1

Supplementary checklist 2

## Data Availability

The datasets analyzed during the current study are all available in the OpenGWAS database (https://gwas.mrcieu.ac.uk/) and MR-base platform (https://www.mrbase.org). Other necessary data were collected from published GWASs (reference 18 and 25). The study can be reproduced according to the methods part step by step.

https://gwas.mrcieu.ac.uk/

## 6.2 Acknowledgements

We appreciate the researchers in the MR-base platform, as well as those who have being collecting and preserving massive GWAS data. We also thank the American Journal Experts (AJE) corporation for their professional native English-Language editing.

## 6 Declarations

### 6.1 Authors’ contributions

Zhongxing Wang takes responsibility for the content of the manuscript. Shaowei Gao and Zhongxing Wang conceived the idea and designed the study. Siyu Luo and Xiaoying Cai searched and reviewed relevant articles. Qiulan He and Chanyan Huang helped collect and assemble data from relevant studies. Shaowei Gao and Huaqiang Zhou did the major work of data extraction and analysis. Writing and edition were provided by all of the authors. The authors meet the criteria for authorship as recommended by the International Committee of Medical Journal Editors (ICMJE). All of the author approved the final manuscript.

### 6.3 Funding

The study was funded by the Key Training Program for Young Teachers of Sun Yat-sen University (19ykzd12) (website: http://www.gzsums.net/news_20459.aspx) and Guangzhou Science and Technology Foundation (201904010418) (website: http://kjj.gz.gov.cn/).

### 6.4 Reporting Checklist

We have completed corresponding checklists: the study was conducted under Burgess’s guidelines (Supplementary checklist 1) and reported according to the STROBE-MR statement (Supplementary checklist 2).

### 6.5 Data Sharing and Data Accessibility

The datasets analyzed during the current study are all available in the OpenGWAS database (https://gwas.mrcieu.ac.uk/) and MR-base platform (https://www.mrbase.org). Other necessary data were collected from the Klimentidis’s and Suri’s published GWASs (reference 18 and 25). The study can be reproduced according to the methods part step by step.

### 6.6 Conflict of Interest

The authors declare that they have no conflict of interest.

### 6.7 Ethical statement

All the data used are summary-level and derived from public genome-wide association studies (GWAS), which had obtained ethical permissions from their respective institutional review boards and written informed consent from their respective participants. Patients and public were not involved in this MR study.

## 6.8 Abbreviations used in the article

CBP: chronic back pain
MR: Mendelian randomization
SNP: single-nucleotide polymorphism
IVW: inverse variance-weighted
GWAS: genome-wide association studies
MR-PRESSO: MR-pleiotropy residual sum and outlier

## Notes

### Competing Interest Statement

The authors have declared no competing interest.

### Clinical Trial

This study is not a clinical trial.

