## Supplementary material for "Investigating the causal relationship between physical activity and chronic back pain: a bidirectional two-sample Mendelian randomization study": Table S1

| Table S1. Characteristics of phenotypes of physical activity for Klimentidis’s GWAS | | | | |
| --- | --- | --- | --- | --- |
| Phenotype | Data type | Mean (SD)/Number of cases | Number of controls | Total number |
| MVPA (MET-minutes/week) | Continuous | 1650 (2084) | - | 337,234 |
| VPA:≥3 vs 0 days/week | Binary | 98,060 | 162,995 | 261,055 |
| SSOE: ≥2-3 vs 0 days/week | Binary | 124,842 | 225,650 | 350,492 |
| Average acceleration (milligravities) | Continuous | 27.98 (27.03) | - | 91,084 |
| Abbreviations: GWAS, genome-wide association studies; MVPA, moderate-vigorous physical activity; VPA, vigorous physical activity; SSOE, strenuous sports or other exercises | | | | |

| Table S2. GWAS-reported SNPs associated with different phenotypes of physical activity for Klimentidis’s GWAS | | | |
| --- | --- | --- | --- |
| SNP ID | Effect allele (alternative) | β (SE) for physical activity | P value |
| MVPA |  |  |  |
| rs2988004 | G (T) | 0.013 (0.002) | 4.1×10^-9^ |
| rs7791992 | A (C) | 0.014 (0.002) | 5.7×10^-10^ |
| rs429358 | C (T) | 0.022 (0.003) | 6.1×10^-13^ |
| rs7804463 | C (T) | -0.015 (0.002) | 1.2×10^-11^ |
| rs3094622 | G (A) | -0.020 (0.003) | 1.4×10^-9^ |
| rs2854277 | T (C) | -0.032 (0.005) | 2.6×10^-10^ |
| rs2035562 | G (A) | 0.014 (0.002) | 3.8×10^-9^ |
| rs149943 | A (G) | -0.019 (0.003) | 2.2×10^-9^ |
| rs1043595 | A (G) | -0.014 (0.002) | 4.3×10^-9^ |
| VPA |  |  |  |
| rs3781411 | T (C) | -0.012 (0.002) | 3.0×10^-10^ |
| rs1248860 | A (G) | 0.010 (0.001) | 1.1×10^-13^ |
| rs2764261 | G (A) | -0.009 (0.001) | 2.0×10^-11^ |
| rs13243553 | A (G) | -0.009 (0.001) | 9.0×10^-11^ |
| rs328902 | T (C) | 0.009 (0.001) | 5.5×10^-10^ |
| SSOE |  |  |  |
| rs159644 | G (A) | 0.007 (0.001) | 1.3×10^-9^ |
| rs166840 | A (G) | -0.008 (0.001) | 3.1×10^-11^ |
| rs10946808 | G (A) | 0.008 (0.001) | 9.9×10^-10^ |
| rs7590676 | C (T) | 0.016 (0.003) | 2.0×10^-9^ |
| rs111901094 | T (G) | -0.009 (0.001) | 3.0×10^-9^ |
| rs62253088 | C (T) | -0.011 (0.001) | 1.0×10^-19^ |
| Average acceleration |  |  |  |
| rs55657917 | G (T) | 0.303 (0.044) | 5.0×10^-12^ |
| rs59499656 | T (A) | 0.228 (0.038) | 2.4×10^-9^ |
| Abbreviations: GWAS, genome-wide association studies; SNP, single-nucleotide polymorphism; MVPA, moderate-vigorous physical activity; VPA, vigorous physical activity; SSOE, strenuous sports or other exercises | | | |

| Table S3. Relaxed-threshold SNPs associated with different phenotypes of physical activity for Klimentidis’s GWAS | | | |
| --- | --- | --- | --- |
| SNP ID | Effect allele (alternative) | β (SE) for physical activity | P value |
| MVPA |  |  |  |
| rs2494664 | G (A) | 0.012 (0.002) | 8.9×10^-8^ |
| rs2942127 | A (G) | -0.016 (0.003) | 3.3×10^-8^ |
| rs10157145 | C (T) | 0.012 (0.002) | 9.6×10^-8^ |
| rs1974771 | A (G) | 0.021 (0.004) | 6.6×10^-9^ |
| rs877483 | C (T) | -0.012 (0.002) | 4.0×10^-8^ |
| rs2035562 | G (A) | 0.014 (0.002) | 3.9×10^-9^ |
| rs2114286 | G (A) | 0.012 (0.002) | 3.3×10^-8^ |
| rs1972763 | T (C) | -0.013 (0.002) | 3.3×10^-8^ |
| rs77742115 | C (T) | 0.018 (0.003) | 9.6×10^-9^ |
| rs114243593 | G (A) | -0.027 (0.005) | 5.5×10^-8^ |
| rs2854277 | T (C) | -0.032 (0.005) | 2.6×10^-10^ |
| rs7804463 | C (T) | -0.015 (0.002) | 1.2×10^-11^ |
| rs1186721 | A (G) | 0.013 (0.002) | 4.4×10^-8^ |
| rs56293069 | C (G) | -0.014 (0.002) | 5.8×10^-8^ |
| rs921915 | G (T) | 0.014 (0.002) | 5.7×10^-10^ |
| rs1043595 | A (G) | -0.014 (0.002) | 4.3×10^-9^ |
| rs67432617 | C (A) | -0.015 (0.003) | 5.1×10^-8^ |
| rs2988004 | G (T) | 0.013 (0.003) | 4.1×10^-9^ |
| rs527737 | T (C) | 0.012 (0.002) | 6.1×10^-8^ |
| rs7326482 | T (G) | 0.013 (0.002) | 1.6×10^-8^ |
| rs10145335 | A (G) | 0.014 (0.002) | 2.7×10^-8^ |
| rs4886868 | G (T) | 0.012 (0.002) | 3.5×10^-8^ |
| rs12912808 | T (C) | -0.018 (0.003) | 1.7×10^-8^ |
| rs429358 | C (T) | 0.022 (0.003) | 6.1×10^-13^ |
| rs1921981 | A (G) | -0.013 (0.002) | 3.8×10^-8^ |
| VPA |  |  |  |
| rs6689056 | A (G) | -0.008 (0.001) | 6.5×10^-8^ |
| rs6667222 | C (A) | -0.009 (0.002) | 8.7×10^-9^ |
| rs1248860 | A (G) | 0.010 (0.001) | 1.1×10^-13^ |
| rs2764261 | G (A) | -0.009 (0.001) | 2.0×10^-11^ |
| rs9276758 | A (G) | -0.008 (0.001) | 1.4×10^-8^ |
| rs13243553 | A (G) | -0.009 (0.001) | 9.0×10^-11^ |
| rs328902 | T (C) | 0.009 (0.001) | 5.5×10^-10^ |
| rs3781411 | T (C) | -0.013 (0.002) | 3.0×10^-10^ |
| rs61866271 | C (G) | -0.028 (0.005) | 6.9×10^-8^ |
| rs58242881 | A (G) | -0.009 (0.002) | 8.6×10^-8^ |
| rs72928932 | G (T) | 0.013 (0.002) | 7.5×10^-8^ |
| rs1461584 | G (A) | 0.009 (0.002) | 8.2×10^-8^ |
| SSOE |  |  |  |
| rs1200154 | A (G) | 0.006 (0.001) | 3.9×10^-8^ |
| rs2994326 | C (T) | 0.008 (0.001) | 4.5×10^-8^ |
| rs288070 | A (G) | 0.011 (0.002) | 1.9×10^-8^ |
| rs6432141 | G (A) | -0.006 (0.001) | 6.9×10^-8^ |
| rs7627864 | G (C) | -0.007 (0.001) | 7.6×10^-9^ |
| rs62253088 | C (T) | -0.011 (0.002) | 1.0×10^-19^ |
| rs4865667 | T (C) | -0.007 (0.001) | 1.0×10^-8^ |
| rs159544 | G (A) | 0.007 (0.001) | 1.3×10^-9^ |
| rs1265178 | A (G) | -0.007 (0.001) | 3.2×10^-8^ |
| rs10946808 | G (A) | 0.008 (0.001) | 9.9×10^-10^ |
| rs896302 | T (C) | -0.007 (0.001) | 1.7×10^-8^ |
| rs551243 | C (G) | -0.006 (0.001) | 6.5×10^-8^ |
| rs4411372 | C (T) | 0.007 (0.001) | 2.0×10^-8^ |
| rs75930676 | C (T) | 0.016 (0.003) | 2.0×10^-9^ |
| rs2470893 | T (C) | -0.006 (0.001) | 5.5×10^-8^ |
| rs166840 | A (G) | -0.008 (0.001) | 3.1×10^-11^ |
| rs111901094 | T (G) | -0.009 (0.001) | 3.0×10^-9^ |
| Average acceleration |  |  |  |
| rs12045968 | G (T) | 0.239 (0.044) | 5.1×10^-8^ |
| rs34517439 | A (C) | -0.308 (0.056) | 4.4×10^-8^ |
| rs6775319 | T (A) | -0.225 (0.041) | 3.5×10^-8^ |
| rs12522261 | A (G) | -0.211 (0.038) | 3.9×10^-8^ |
| rs9293503 | C (T) | -0.329 (0.059) | 2.1×10^-8^ |
| rs11012732 | G (A) | -0.225 (0.039) | 5.4×10^-9^ |
| rs148193266 | C (A) | 0.510 (0.092) | 3.1×10^-8^ |
| rs1550435 | C (T) | -0.200 (0.037) | 5.0×10^-8^ |
| rs56194509 | G (T) | 0.303 (0.043) | 5.0×10^-12^ |
| rs59499656 | T (A) | 0.228 (0.038) | 2.4×10^-9^ |
| Abbreviations: GWAS, genome-wide association studies; SNP, single-nucleotide polymorphism; MVPA, moderate-vigorous physical activity; VPA, vigorous physical activity; SSOE, strenuous sports or other exercises | | | |

| Table S4. SNPs associated with chronic back pain reported by Suri’s genome-wide meta-analysis | | | | |
| --- | --- | --- | --- | --- |
| SNP ID | Effect allele (alternative) | OR (SE) for CBP | β (SE) for CBP | P value |
| rs12310519 | T (C) | 1.07 (0.008) | 0.068 (0.010) | 4.5×10^-19^ |
| rs1453867 | C (T) | 0.97 (0.006) | -0.030 (0.005) | 3.9×10^-7^ |
| rs4384683 | G (A) | 0.97 (0.006) | -0.030 (0.005) | 2.4×10^-10^ |
| rs7833174 | T (C) | 1.05 (0.007) | 0.049 (0.005) | 4.4×10^-13^ |
| Abbreviations: SNP, single-nucleotide polymorphism; OR, odds ratio; CBP, chronic back pain | | | | |

| Table S5. Cohorts of chronic back pain used as outcome cohorts | | | | |
| --- | --- | --- | --- | --- |
| Name of dataset | Population | Number of cases | Number of controls | Number of SNPs |
| ukb-a-346 | European | 57,893 | 26,596 | 10,894,596 |
| ukb-b-8463 | European | 80,588 | 36,816 | 9,851,867 |
| ukb-e-3571_CSA | South Asia | 1883 | 916 | 9,810,007 |
| ukb-e-3571_AFR | African American or Afro-Caribbean | 1352 | 675 | 15,478,660 |
| Abbreviations: SNP, single-nucleotide polymorphism | | | | |

| Table S6. Matrix of pooled IVW effect sizes of different phenotypes of physical activity on different cohorts of chronic back pain | | | | |
| --- | --- | --- | --- | --- |
| OR (95% CI) | ukb-a-346 | ukb-b-8463 | ukb-e-3571_CSA | ukb-e-3571_AFR |
| GWAS-reported |  |  |  |  |
| MVPA | 0.95 (0.82-1.10) | 0.98 (0.85-1.13) | 0.63 (0.03-11.47) | 4.26 (0.01-1.6E03) |
| VPA | 0.88 (0.69-1.11) | 0.92 (0.74-1.15) | 1.88 (0.005-7.42) | 8.63 (2.2-1.4E04)* |
| SSOE | 0.81 (0.52-1.27) | 0.85 (0.58-1.25) | 347 (0.88-14E05) | 3.3E03 (0.64-1.7E07) |
| Average acceleration | 1.0 (0.98-1.03) | 1.0 (0.98-1.02) | 0.69 (0.41-1.19) | 0.74 (0.40-1.36) |
| Relaxed threshold |  |  |  |  |
| MVPA | 1.06 (0.96-1.17) | 1.06 (0.97-1.16) | 0.74 (0.11-4.85) | 12 (0.86-172) |
| VPA | 0.89 (0.71, 1.10) | 0.93 (0.76-1.14) | 2.67 (0.03-257) | 221 (0.75-6.6E04) |
| SSOE | 0.82 (0.66-1.02) | 0.84 (0.70-1.02) | 5.93 (0.02-2038) | 3.10 (0.01-1108) |
| Average acceleration | 1.0 (0.99-1.01) | 1.0 (0.99-1.00) | 0.91 (0.72-1.16) | 0.84 (0.63-1.13) |
| ^*^ p < 0.05  Abbreviations: IVW, inverse variance weighted; OR, odds ratio; CI, confidence interval; GWAS, genome-wide association studies; MVPA, moderate-vigorous physical activity; VPA, vigorous physical activity; SSOE, strenuous sports or other exercises | | | | |

Figure S1

Plots of sensitivity analyses for moderate-vigorous physical activity on chronic back pain. (A) Leave-one-out analysis. Each point and corresponding black line indicate the pooled effect size and corresponding standard error, respectively, of all SNPs except the one marked on the left. (B) Funnel plot of 1/standard error against the corresponding effect size (β) for all SNPs: each point refers to an individual SNP; the vertical line indicates the pooled effect size with the inverse variance-weighted method.

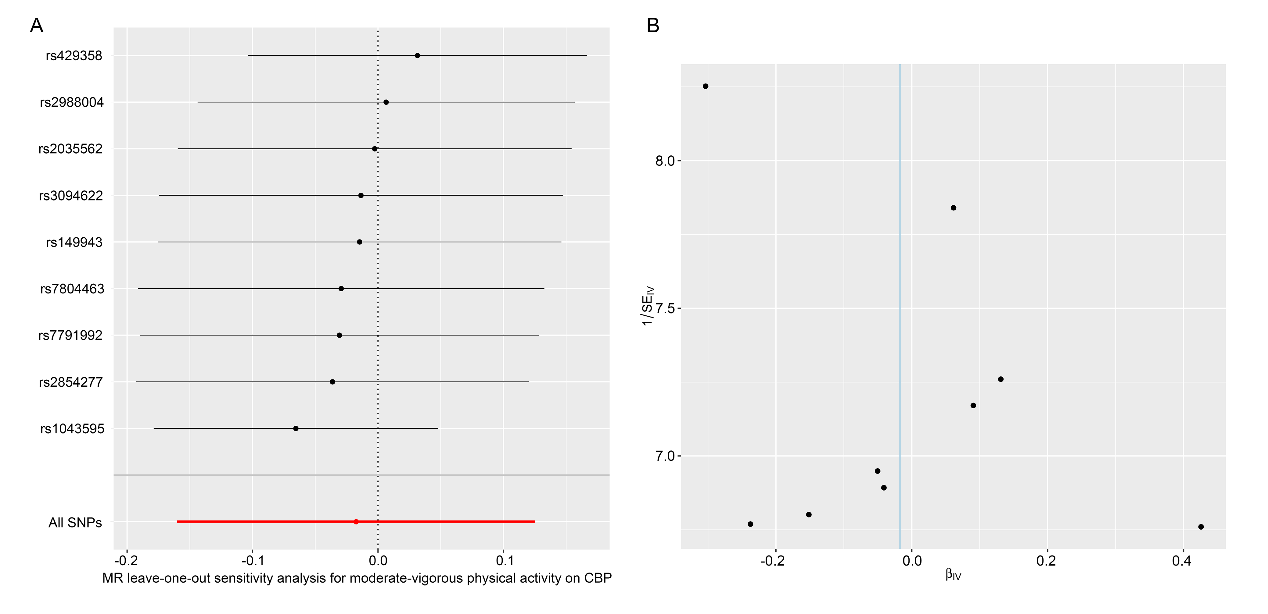

| Table S7. Matrix of pooled IVW effect sizes of chronic back pain on different phenotypes of physical activity | | | | |
| --- | --- | --- | --- | --- |
| β (95% CI) | MVPA | VPA | SSOE | Average acceleration |
| All SNPs for CBP | -0.07 (-0.12, -0.01)* | -0.05 (-0.09, -0.01)* | -0.04 (-0.06, -0.02)* | -1.28 (-2.20, -0.36)* |
| All SNPs for CBP except rs1453867 | -0.07 (-0.13, -0.01)* | -0.05 (-0.09, -0.01)* | -0.04 (-0.07, -0.01)* | -1.43 (-2.51, -0.25)* |
| * p < 0.05  Abbreviations: SNP, single-nucleotide polymorphism; CBP, chronic back pain; MVPA, moderate-vigorous physical activity; VPA, vigorous physical activity; SSOE, strenuous sports or other exercises | | | | |

Figure S2.

Plots of sensitivity analyses for chronic back pain on moderate-vigorous physical activity. (A) Leave-one-out analysis. Each point and corresponding black line indicate the pooled effect size and standard error, respectively, of all SNPs except the one marked on the left. (B) Funnel plot of the 1/standard error against corresponding effect size (β) for all SNPs: each point refers to an individual SNP; the vertical line indicates the pooled effect size with the inverse variance-weighted method.

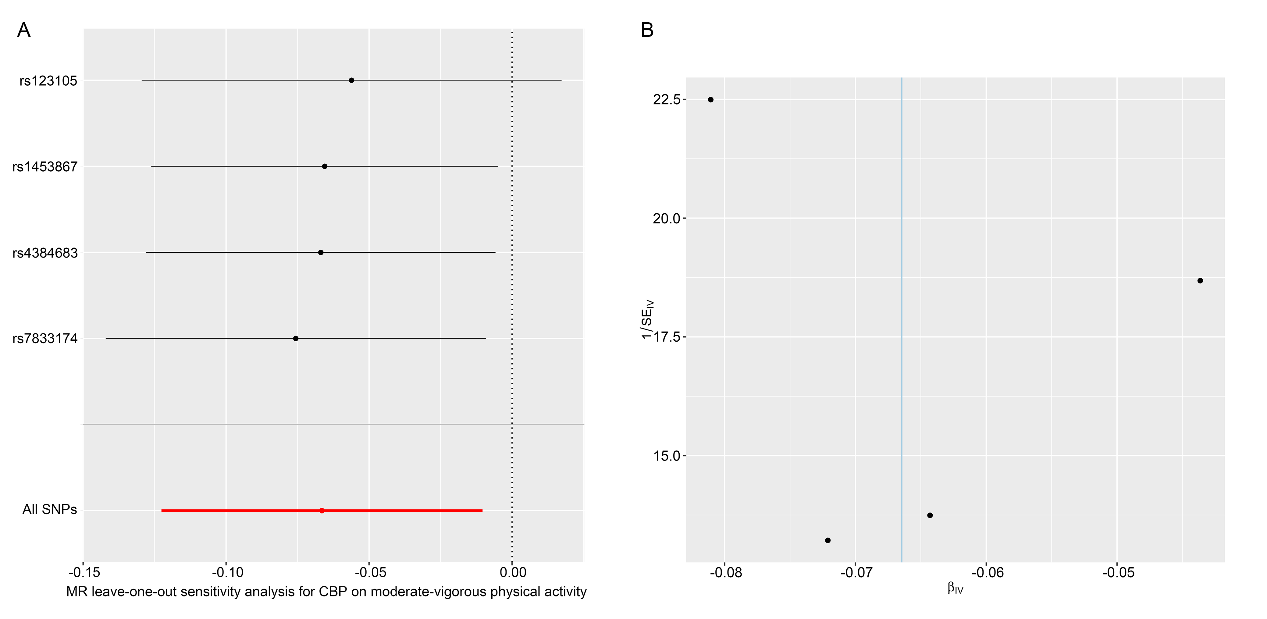

| Table S8. Manually detected potential pleiotropy in the Phenoscanner database | | | | |
| --- | --- | --- | --- | --- |
| SNP | Potential pleiotropic trait | Effect allele (Alternate allele) | Beta | P value |
| Physical activity to CBP |  |  |  |  |
| rs1043595 | - |  |  |  |
| rs2854277 | Rheumatoid arthritis; | T (C) | -0.4463 | 8.90×10^-32^ |
|  | Trunk fat-free mass | A (C) | 0.02448 | 6.94×10^-30^ |
| rs7791992 | - |  |  |  |
| rs7804463 | - |  |  |  |
| rs149943 | Trunk fat mass | C (T) | 0.01889 | 3.57×10^-8^ |
| rs3094622 | Trunk fat-free mass | G (A) | 0.01396 | 3.14×10^-10^ |
| rs2035562 | Trunk fat-free mass | A (G) | -0.0115 | 9.18×10^-12^ |
| rs2988004 | - |  |  |  |
| rs429358 | Trunk fat mass | C (T) | -0.02184 | 8.91×10^-11^ |
| CBP to activity |  |  |  |  |
| rs7833174 | Trunk fat-free mass | C (T) | -0.01802 | 2.89×10^-23^ |
| rs4384683 | - |  |  |  |
| rs1453867 | Trunk fat-free mass | T (G) | 0.009068 | 6.75×10^-9^ |
| rs12310519 | - |  |  |  |
| Abbreviations: CBP, chronic back pain | | | | |

Figure S3

Forest plots of individual and pooled MR effect sizes for the bidirectional association between moderate-vigorous physical activity and CBP after removing SNPs with potential pleiotropy. (A) Forest plot for the direction from moderate-vigorous physical activity to CBP. (B) Forest plot for the direction from CBP to moderate-vigorous physical activity.

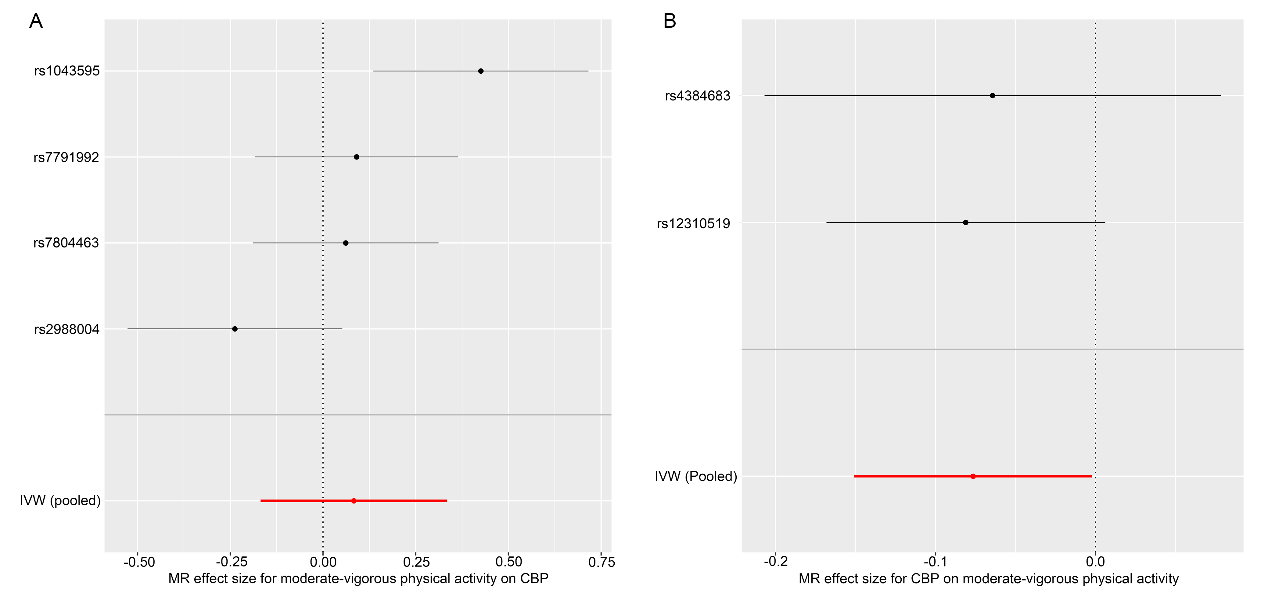

Figure S4

Plots for leave-one-out analysis of chronic back pain on the three other phenotypes of physical activity: (A) vigorous physical activity; (B) strenuous sports or other exercise; and (C) acceleration-based physical activity. Each point and corresponding black line indicate the pooled effect size and standard error, respectively, of all SNPs except the one marked on the left.

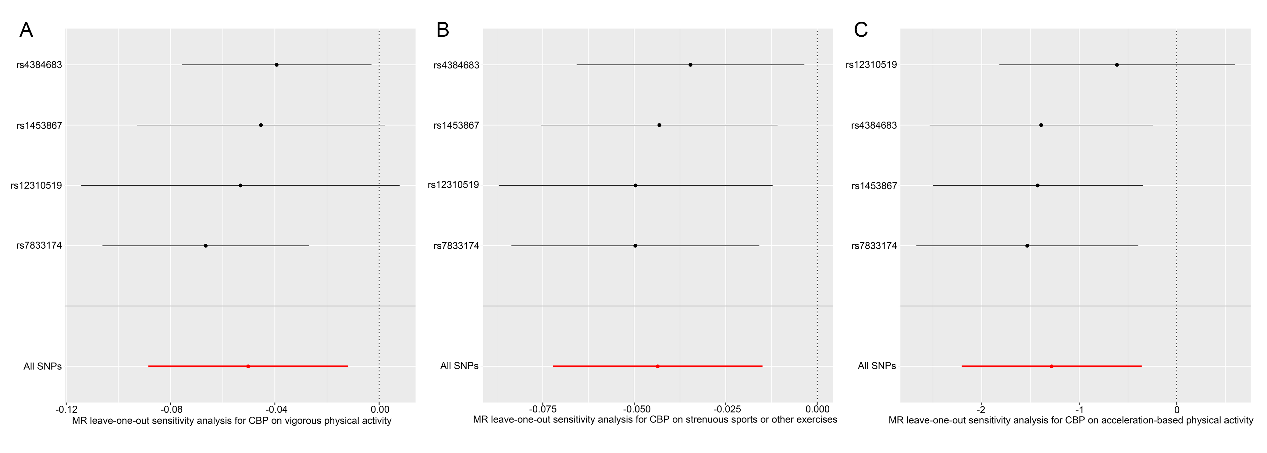
