## Supplementary checklist 1 for "Investigating the causal relationship between physical activity and chronic back pain: a bidirectional two-sample Mendelian randomization study"

**Checklist for reviewing Mendelian randomization investigations, recommended by Burgess’s MR guidelines^1^.**

1. **What is the primary hypothesis of interest? What is the motivation for using Mendelian randomization? What is the scope of the investigation? What and how many primary analyses?**

The primary hypothesis of interest is that physical activity has inverse causal association with chronic back pain (page 3). The motivation to use mendelian randomization is that even though recent studies indicates a negative association between physical activity and chronic back pain, the studies with high-level evidence (like randomized control study), which can address the problem of causal inference, are lacking (page 3). The scope of the investigation is to understand aetiology for a symptom. The primary analysis is the bidirectional mendelian randomization between self-reported moderate-vigorous physical activity and self-reported chronic back pain (page 3-4).

2. **(Data sources) What type of mendelian randomization investigation is this? One-sample or two-sample? Sample overlap? Summarized data or individual-level data? Drawn from same population? Relevence to applied research?**

This study is a two-sample mendelian randomization investigation, using summarized data (page 3). The data sources for exposure and outcome were drawn from same population, having sample overlap which is hard to quantitatively estimated. The relevant research for physical activity is Klimentidis’s GWAS (reference 18 in the article), for chronic back pain is Suri’s genome wide meta-analysis (reference 25 in the article).

3. **Selection of genetic variants – how were the genetic variants chosen? Single or multiple gene regions?**
a. Biological rationale?
b. GWAS analysis? If so, what dataset? What was the p-value threshold? Clumping
c. Were genetic variants excluded from the analysis? Associations with pleiotropic pathways?
d. How else was the validity of genetic variants as instrumental variables assessed?

We chose variants for multiple gene regions, using summarized results of GWAS (page 3). The P-value is less than 5×10^-8^ primarily, using clumping (page 4). We excluded some variants when conduct sensitivity analyses, with the method of MR-PRESSO. The validity of genetic variants was assessed by the heritability calculated in the relevant GWAS (page 3-5).

4. **Variant harmonization: was it checked that genetic variants were appropriately orientated across the datasets.**

Yes, we checked the effect allele and its frequency of each instrument, which is required to determine the direction of strand (page 4).

5. **(Primary analysis) What was the primary analysis? What was the statistical method? How implemented? Multiple testing?**

The primary analysis is the bidirectional mendelian randomization between self-reported moderate-vigorous physical activity and self-reported chronic back pain with the method of multiplicative random-effect inverse variance-weighted meta-analysis. There is multiple testing, using different sub-phenotypes and cohorts (Page 3-5).

**6 and 7. (Supplementary and sensitivity analyses) What analyses were performed to support and assess the validity of the primary analysis?**

A series of methods were applied for sensitivity analyses: in addition to setting multiple comparisons among different phenotypes and different cohorts, alternative statistical methods including the weighted median method and MR egger regression, the funnel plot, Cochran’s Q statistic, leave-one-out analyses, MR-PRESSO (Pleiotropy Residual Sum and Outlier), and the MR Egger intercept test of deviation from the null were used for detecting heterogeneity and horizontal pleiotropy, which is critical to support and assess the validity of the primary analysis (Page 5).

**8. (Data presentation) How are the data and results presented to allow readers to assess the analysis and assumption?**

We used scatter plot, forest plot, funnel plot, leave-one-out analysis plot, and comparison of different methods (page 5-6).

**9. (Interpretation) How have results been interpreted, particularly any numerical estimates?**

From the bidirectional mendelian randomization study, we found the causal inference in the direction of chronic back pain to physical activity, but not in the inverse direction. We also reported the numerical estimates, but it was just for a reference, never over-interpreted (page 5-6).
