## Supplementary checklist 2 for "Investigating the causal relationship between physical activity and chronic back pain: a bidirectional two-sample Mendelian randomization study"

**STROBE-MR: Guidelines for strengthening the reporting of Mendelian randomization studies[1]**

1. TITLE and ABSTRACT

Indicate Mendelian randomization as the study’s design in the title and/or the abstract.

Yes, at page 1.

INTRODUCTION

2. Background

Explain the scientific background and rationale for the reported study. Is causality between exposure and outcome plausible? Justify why MR is a helpful method to address the study question.

Yes, at page 2-3.

3. Objectives

State specific objectives clearly, including pre-specified causal hypotheses (if any).

Yes, at page 3.

METHODS

4. Study design and data sources

Present key elements of study design early in the paper. Consider including a table listing sources of data for all phases of the study. For each data source contributing to the analysis, describe the following:

a) Describe the study design and the underlying population from which it was drawn. Describe also the setting, locations, and relevant dates, including periods of recruitment, exposure, follow-up, and data collection, if available.

b) Give the eligibility criteria, and the sources and methods of selection of participants.

c) Explain how the analyzed sample size was arrived at.

d) Describe measurement, quality and selection of genetic variants.

e) For each exposure, outcome and other relevant variables, describe methods of assessment

and, in the case of diseases, the diagnostic criteria used.

f) Provide details of ethics committee approval and participant informed consent, if relevant.

Yes, we have included a table listing data sources (Table S1-S5). The description of all above is present in page 3-5.

5. Assumptions

Explicitly state assumptions for the main analysis (e.g. relevance, exclusion, independence, homogeneity) as well assumptions for any additional or sensitivity analysis.

Yes, at page 3-5.

6. Statistical methods: main analysis

Describe statistical methods and statistics used.

a) Describe how quantitative variables were handled in the analyses (i.e., scale, units, model).

b) Describe the process for identifying genetic variants and weights to be included in the analyses (i.e, independence and model). Consider a flow diagram.

c) Describe the MR estimator, e.g. two-stage least squares, Wald ratio, and related statistics. Detail the included covariates and, in case of two-sample MR, whether the same covariate set was used for adjustment in the two samples.

d) Explain how missing data were addressed.

e) If applicable, say how multiple testing was dealt with.

Yes, at page 3-5.

7. Assessment of assumptions

Describe any methods used to assess the assumptions or justify their validity.

Yes, at page 4-5.

8. Sensitivity analyses

Describe any sensitivity analyses or additional analyses performed.

Yes, at page 5.

9. Software and pre-registration

a) Name statistical software and package(s), including version and settings used.

b) State whether the study protocol and details were pre-registered (as well as when and where).

Yes, name of statistical software and packages are declared at page 4. The study protocol has not been pre-registered or published.

RESULTS

10. Descriptive data

a) Report the numbers of individuals at each stage of included studies and reasons for exclusion. Consider use of a flow-diagram. Yes, but the numbers of individuals have been reported at the Methods part. The flow-diagram is present.

b) Report summary statistics for phenotypic exposure(s), outcome(s) and other relevant variables (e.g. means, standard deviations, proportions). Yes, the summary statistics of phenotypic exposure(s), outcome(s) and other relevant variables are in Table S1-S5.

c) If the data sources include meta-analyses of previous studies, provide the number of studies, their reported ancestry, if available, and assessments of heterogeneity across these studies. Consider using a supplementary table for each data source. Yes, at page 4.

d) For two-sample Mendelian randomization:

i. Provide information on the similarity of the genetic variant-exposure associations between the exposure and outcome samples. Yes, the scatter plot can provide this information.

ii. Provide information on extent of sample overlap between the exposure and outcome data sources. We reported there is sample overlap but the extent is hard to quantitatively estimated

11. Main results

a) Report the associations between genetic variant and exposure, and between genetic variant and outcome, preferably on an interpretable scale (e.g. comparing 25th and 75^th^ percentile of allele count or genetic risk score, if individual-level data available). This study is a two-sample MR study without individual-level data.

b) Report causal effect estimate between exposure and outcome, and the measures of uncertainty from the MR analysis. Use an intuitive scale, such as odds ratio, or relative risk, per standard deviation difference. Yes, at page 5-6.

c) If relevant, consider translating estimates of relative risk into absolute risk for a meaningful time-period. Yes, at page 5-6.

d) Consider any plots to visualize results (e.g. forest plot, scatterplot of associations between

genetic variants and outcome versus between genetic variants and exposure). Yes, at page 5-6.

12. Assessment of assumptions

a) Assess the validity of the assumptions.

b) Report any additional statistics (e.g., assessments of heterogeneity, such as I2, Q statistic).

Yes, at page 5-6.

13. Sensitivity and additional analyses

a) Use sensitivity analyses to assess the robustness of the main results to violations of the

assumptions. Yes, at page 5-6.

b) Report results from other sensitivity analyses (e.g., replication study with different dataset, analyses of subgroups, validation of instrument(s), simulations, etc.). Yes, at page 5-6.

c) Report any assessment of direction of causality (e.g., bidirectional MR). Yes.

d) When relevant, report and compare with estimates from non-MR analyses. Yes, at page 6.

e) Consider any additional plots to visualize results (e.g., leave-one-out analyses). Yes, at page 5-6.

DISCUSSION

14. Key results

Summarize key results with reference to study objectives.
Yes, at page 6.

15. Limitations

Discuss limitations of the study, taking into account the validity of the MR assumptions, other sources of potential bias, and imprecision. Discuss both direction and magnitude of any potential bias, and any efforts to address them.

Yes, at page 7

16. Interpretation

a) Give a cautious overall interpretation of results considering objectives and limitations. Compare with results from other relevant studies.

b) Discuss underlying biological mechanisms that could be modelled by using the genetic variants to assess the relationship between the exposure and the outcome.

c) Discuss whether the results have clinical or policy relevance, and whether interventions could have the same size effect.

Yes, at page 6-8.

17. Generalizability

Discuss the generalizability of the study results (a) to other populations (i.e. external validity), (b) across other exposure periods/timings, and (c) across other levels of exposure.

Yes, we used four cohort of chronic back pain from different population; we also used four sub-phenotypes of physical activity.

OTHER INFORMATION

18. Funding

Give the source of funding and the role of the funders for the present study and, if applicable, for

the original study or studies on which the present article is based.

Yes, at page 8.

19. Data and data sharing

Present data used to perform all analyses or report where and how the data can be accessed. State

whether statistical code is publicly accessible and if so, where.

Yes, at page 8-9.

20. Conflicts of Interest

All authors should declare all potential conflicts of interest.

Yes, at page 9.
